# Keep calm and carry on: safety, feasibility and early outcomes of head and neck cancer treatment during the COVID-19 pandemic

**DOI:** 10.1101/2020.08.18.20167270

**Authors:** Sara Walker, Maureen Thomson, Frances Campbell, Lisa Hay, Derek Grose, Allan James, Carolynn Lamb, Ioanna Nixon, Stefano Schipani, Christina Wilson, Claire Paterson

## Abstract

**Background:** Patients with cancer are considered at higher risk of COVID-19 infection and increased severity of infection. Anti-cancer treatment may further increase those risks.

The aim of this work is to report early outcomes in patients with head and neck cancer (HNC) treated during the pandemic.

**Materials and Methods:** A retrospective cohort study in a UK tertiary level oncology centre between 1^st^ March and 23 June 2020, including patients with HNC who were either newly diagnosed, had developed new recurrent/metastatic disease, or were already scheduled to receive treatment during that period.

**Results:** 200 patients were evaluated. Median age was 64 years, 65.5% had multiple co-morbidities, 77.5% were current or ex-smokers and 59.5% lived in areas of deprivation. 99 patients were treated with 6 weeks of radical (chemo) radiotherapy. Systemic anti-cancer treatment was delivered to 40 patients.

2 (1.0%) patients with HNC had confirmed COVID-19 infection; 1 patient prior to primary radical RT - no delay to treatment was required and RT was completed as planned, 1 patient acquired COVID-19 after primary surgery but recovered well and started adjuvant RT 9.7 weeks after surgery.

The proportion of patients receiving supportive care only (19.5%) was in keeping with that pre-COVID-19. The proportion of patients not completing (chemo) radiotherapy (3.4%) or with gaps in treatment (14.1%) was similar to pre-COVID-19. 30-day mortality after radical (chemo)radiotherapy was 2.3%, no higher than in previous years.

**Conclusions:** It is feasible and safe to deliver standard treatment for patients with HNC during the COVID-19 pandemic.

## Introduction

The World Health Organisation (WHO) declared the corona virus disease-19 (COVID-19) outbreak a pandemic on 11 March 2020. [1] The index case in Scotland had already been confirmed on the 1st March 2020. [2] To date, there have been 18781 laboratory confirmed cases in Scotland and of these, 2491 people have died. [3] The west of Scotland (WoS) managed clinical network (MCN) for head and neck cancer (HNC) serves a population of 2.5 million people in four NHS Boards. [4] Of these NHS Greater Glasgow and Clyde has recorded the highest number of COVID-19 cases with 4983 people testing positive and an infection rate of 416 cases per 100 000 population. [3] The WoS MCN records, on average, 630 cases of HNC annually. [4-7] It is well recognized that our cancer network covers some of the most deprived areas in Scotland [8] and this is associated with a disproportionate burden of co-morbidity. [9] As early data emerged from China and Italy, there was concern that patients with cancer may represent a group particularly vulnerable to COVID-19 with higher incidence, severity and mortality compared to the general population. [10 - 12] Cautions were issued about the use of anti-cancer treatments central to the management of HNC such as cytotoxic chemotherapy, immunotherapy, radiotherapy (RT) and surgery. [13-15] Equally, the risk of delaying cancer care was acknowledged with patients at risk of disease progression and death while waiting for treatment. [13] Other co-morbidities, such as cardiovascular disease, diabetes and chronic pulmonary disease, highly prevalent in our HNC population, as well as deprivation, were noted to be risk factors for severity of COVID-19 infection. [16 - 18] Our HNC multi-disciplinary team (MDT) faced a significant challenge; achieving a balance between adequate and timely treatment of cancer in the face of the risk to our patients from this potentially deadly infection. [19] The aim of this work is, for the first time, to describe the early outcomes of patients with HNC treated during the peak of the outbreak, demonstrating that it is feasible and safe to deliver anti-cancer treatment for HNC despite the ongoing COVID-19 pandemic.

## Materials and Methods

This was a retrospective cohort study in a UK tertiary level oncology centre.

### Study population

Eligible patients were those in our cancer network with a diagnosis of HNC who were either newly diagnosed, had developed new recurrent or metastatic disease, or had previously been diagnosed and were scheduled to receive treatment between 1^st^ March – 23^rd^ June 2020.

### Data collection and analysis

Patients who were diagnosed, either de novo or with new recurrent/metastatic disease were identified from the database generated by regional tumour board meetings. Patients scheduled to receive either RT or systemic anti-cancer treatments (SACT) were identified from the RT bookings system and the electronic prescribing system respectively. The electronic patient record was reviewed for each case. Patients’ social deprivation quintile was determined using the Scottish Index of Multiple Deprivation (SIMD). [20] The West of Scotland research ethics service waived ethical approval for this evaluation of our standard of care service. Data was stored securely and accessed only by the clinical team. Descriptive statistical analysis was performed.

### Treatment

Standard treatment regimens and any modifications to them are described in table 1. Concurrent cisplatin is used for patients with locally advanced HNSCC, aged 70 years or less and with no contraindications. At least 140mg/m^2^ is required to provide a survival advantage. While modelling from phase III trials show a progressive improvement in survival with 10 mg/m^2^ cisplatin increments above 140mg/m^2^, it is unclear if this advantage is due to the higher dose itself or simply fitter patients tolerating additional cisplatin, hence our temporary dose reduction to 75mg/m^2^ on day 1 and 22 of RT. [21]

**Table 1.** Treatment regimens and modifications as a result of COVID-19 pandemic

| Treatment modality | Treatment intent | Regimen | Modified during pandemic |
| --- | --- | --- | --- |
| Radiotherapy | Primary radical | 65Gy/30#<br>70Gy/35# (re-irradiation or close to neural structures)<br>55Gy/20# (early stage larynx ca, primary site only) | No |
|  | Adjuvant to surgery | 65Gy/30# (+ve resection margins or ENE)<br>60Gy/30# | No |
|  | Palliative | 50Gy/25#, 40Gy/15#, 20Gy/5#, single 8Gy#<br>Depending on disease burden/patient fitness | No |
| SACT | Induction | Cisplatin 75mg/m<sup>2</sup> & 5FU 750mg/m<sup>2</sup> d1-4 via PICC line q21 | No |
|  |  | Carboplatin AUC5 & Paclitaxel 175mg/m<sup>2</sup> q21 | No |
|  | Concurrent | Cisplatin 100mg/m<sup>2</sup> d1 & 22 of RT | Yes, reduced to 75mg/m<sup>2</sup> d1 & 22 |
|  |  | Cetuximab 400mg/m<sup>2</sup> loading dose week prior to RT, 250mg/m<sup>2</sup> weekly during RT | No |
|  | Palliative | Carboplatin AUC5 & Paclitaxel 175mg/m<sup>2</sup> q21 | No |
|  |  | Cisplatin 75mg/m<sup>2</sup> or Carboplatin AUC5 & 5FU 750mg/m<sup>2</sup> d1-4 q21 | No |
|  |  | Cisplatin 75mg/m<sup>2</sup> or Carboplatin AUC5 & capecitabine 625mg/m<sup>2</sup> bd q21 | No |
|  |  | Nivolumab 240mg q14 (2<sup>nd</sup> line) | Yes, option to use nivolumab 480mg q28 2-3 months after initiation of treatment [23] |
|  |  | Pembrolizumab 200mg q21 (1<sup>st</sup> line) | New treatment option, not available prior to pandemic |

Concurrent cetuximab is used for patients with specific contra-indications to cisplatin in the primary setting. [23] Three cycles of induction chemotherapy is standardly given for nasopharyngeal cancer. During the pandemic, consideration was given on a case-by-case basis to delivering 2 instead of 3 cycles in view of data suggesting this may provide equivalent benefit. [24]

### Practical measures for risk mitigation

While patients with a new diagnosis of HNC were reviewed face to face in clinic, assessment prior to each cycle of SACT was carried out by telephone consultation. On-treatment reviews to manage toxicity for patients undergoing radical RT or CRT was done by telephone in the first 3 weeks of treatment; patients in the latter stages of treatment were assessed face to face.

All patients attending our cancer centre are screened for COVID-19 symptoms on entry. Symptomatic patients are placed in an isolation room and COVID-19 testing carried out. A dedicated linear accelerator is available to treat patients with COVID-19. In-patients are tested for COVID-19 on admission. Tests are repeated every 4 days while the patient remains in hospital. A dedicated ‘COVID-19 ward’ separate to the cancer centre accommodates any patients with confirmed COVID-19.

Patients undergoing or completing anti-cancer treatment are advised to ‘shield’. [25]

Patients attend clinic appointments on their own and in-patient visiting has been suspended. Waiting and treatment areas have been reconfigured to comply with 2m social distancing policy. [26] All clinical staff wear personal protective equipment (PPE) when in contact with all patients, regardless of COVID-19 status. [27] On-line videoconferences have replaced face-to-face multi-disciplinary team meetings.

### Confirmation of COVID-19 infection

Patients considered to have COVID-19 infection are those with a positive polymerase chain reaction (PCR) test from throat or nasopharyngeal swabs for 2019 NCoV. In keeping with WHO guidance, appearances on chest imaging alone were not used to make a diagnosis of COVID-19. [28] Patients were not routinely tested for COVID-19 prior to embarking on SACT or RT.

## Results

200 patients were evaluated – 101 with a new diagnosis of HNC, 19 with recurrent disease and 80 who had been diagnosed prior to the pandemic and were scheduled to receive treatment in the period under consideration. Demographics for the whole cohort are shown in table 2. Only 2 patients (1.0%) had confirmed COVID-19 infection during the course of their diagnosis or treatment for HNC.

**Table 2.** Patient demographics & tumour details

|  | Intent of Treatment |  |  |  |
| --- | --- | --- | --- | --- |
|  | Number (% of group) |  |  |  |
|  | Whole cohort<br>200 (100%) | Radical Treatment<br>117 (58.5%) | Palliative Treatment<br>44 (22%) | Palliative Care<br>39 (19.5%) |
| **<u>Gender</u>** |  |  |  |  |
| Female | 51 (25.5%) | 29 (24.8%) | 8 (18.2%) | 14 (35.9%) |
| Male | 149 (74.5%) | 88 (75.2%) | 36 (81.8%) | 25 (64.1%) |
| **Median age, years (range)** | 64 (30-95) | 62 (30-86) | 59 (37-95) | 71(40-91) |
| **Presence of multiple co-morbidities** | 131 (65.5%) | 76 (65.0%) | 25 (56.8%) | 30 (76.9%) |
| **<u>WHO Performance status</u>** |  |  |  |  |
| 0 | 87 (43.5%) | 74 (63.2%) | 8 (18.2%) | 5 (12.8%) |
| 1 | 77 (38.5%) | 36 (30.8%) | 34 (77.3%) | 7 (18.0%) |
| 2 | 23 (11.5%) | 7 (6.0%) | 2 (4.5%) | 14 (35.9%) |
| 3+ | 13 (6.5%) | 0 | 0 | 13 (33.3%) |
| **<u>Smoking status</u>** |  |  |  |  |
| Current or former smoker | 155 (77.5%) | 88 (75.2%) | 37 (84.1%) | 30 (76.9%) |
| Non-smoker | 45 (22.5%) | 29 (24.8%) | 7 (15.9%) | 9 (23.1%) |
| **<u>SIMD quintile</u>** |  |  |  |  |
| 1 | 78 (39%) | 45 (38.5%) | 18 (40.9%) | 15 (38.5%) |
| 2 | 41 (20.5%) | 28 (23.9%) | 7 (15.9%) | 6 (15.4%) |
| 3 | 33 (16.5%) | 14 (12.0%) | 6 (13.6%) | 13 (33.3%) |
| 4 | 25 (12.5%) | 17 (14.5%) | 5 (11.4%) | 3 (7.7%) |
| 5 | 23 (11.5%) | 13 (11.1%) | 8 (18.2%) | 2 (5.1%) |
| **<u>Histological sub-type</u>** |  |  |  |  |
| SCC | 174 (87.0%) | 102 (87.2%) | 39 (88.6%) | 33 (84.6%) |
| Non-SCC | 26 (13.0%) | 15 (12.8%) | 5 (11.4%) | 6 (15.4%) |

### Radical treatments

117 patients received radical treatment during this period as described in table 3.

**Table 3.** Number of courses of treatment delivered by treatment type

| Treatment |  |  | Courses of Treatment<br>n= 201 (%) |
| --- | --- | --- | --- |
| Radical<br>n=117 | Surgery alone |  | 17 (8.5) |
|  | Primary RT |  | 40 (20.0) |
|  | Induction<br>chemotherapy<br>followed by | Radical RT | 6 (3) |
|  |  | CRT | 1 (0.5) |
|  |  | Unknown | 1 (0.5)* |
|  | Primary chemoRT |  | 30 (15.0) |
|  | Cetuximab-RT |  | 2 (1.0) |
|  | Adjuvant RT |  | 17 (8.5) |
|  | Adjuvant CRT |  | 3 (1.5) |
| Palliative<br>n= 84 | Radiotherapy |  | 13 (6.5) ** |
|  | SACT |  | 32 (16) ** |
|  | Palliative care |  | 39 (19.5) |

\*patient relocated out with cancer network
\*\*1 patient received both SACT and palliative RT

17 (8.5%) patients underwent surgery alone with no adjuvant treatment; this was not modified on the basis of COVID-19. No patient received RT or CRT as an alternative to surgery.

Of 54 patients with stage III or IV squamous cell cancer (SCC) receiving non-surgical primary treatment only 31 of 54 (57.4%) received concurrent CRT. However, the decision to deliver RT alone in the remainder was for reasons other than risk of COVID-19 infection in all but 3 patients (co-morbidities, age >70 years, T3N0 disease and re-irradiation). 31 patients received primary radical CRT for HNC. 7 patients are yet to reach day 22 of the schedule so have been censored. Mean dose density achieved for evaluable patients (23) was 151mg/m^2^ (range 75mg/m^2^ -200mg/m^2^). Reasons for dose reduction beyond that previously described were treatment related toxicities (4), deranged LFTS (1), renal dysfunction (1), parotiditis (1).

2 patients received cetuximab-RT. 1 patient received all planned cetuximab infusions. 1 patient received only 5 of 7 cetuximab infusions due to treatment related toxicities.

Of the 46 patients who received primary radical RT, 1 patient had confirmed COVID-19 infection prior to starting treatment. No delay or modification to treatment delivery was required and RT was completed as planned. 20 patients received adjuvant treatment; 17 RT alone and 3 CRT. No patient with high risk pathological features post-resection (positive margin and/or extra nodal extension) was denied CRT because of the COVID-19 pandemic. However, only 1 patient completed both cycles of concurrent chemotherapy as planned. The 2^nd^ cycle of cisplatin was omitted in 2 patients; 1 because of chemotherapy induced toxicity and 1 because of COVID-19 risk. 1 patient who developed COVID-19 in the post-operative period received adjuvant RT 9.7 weeks after surgery and completed it as scheduled.

The proportion of patients who completed (C)RT as primary or adjuvant treatment as well as the frequency of gaps in treatment are shown in table 4. 88 patients have completed treatment and 30 day follow up, the remaining have been censored. Three courses of RT were discontinued earlier than planned; 1 because of toxicities and 2 due to sudden unexpected deaths in the early stages of RT in patients with no preceding illness. 30-day mortality rate of patients receiving radical (primary or adjuvant) RT or CRT was 3.3% (2 of 61) and 0% (0 of 27) respectively.

**Table 4.** Proportion patients completing treatment and frequency of gaps in treatment.

| Time period |  | Number of patients (%) |  |  |  |  |  |
| --- | --- | --- | --- | --- | --- | --- | --- |
|  |  | Total | Completed RT course | Duration of gaps in RT<br>(of % of those who completed treatment) |  |  |  |
|  |  |  |  | Any gap | 1 day | 2 days | 3+ days |
| COVID-19<br>Pandemic (1<sup>st</sup><br>March -23<sup>rd</sup><br>June 2020) | RT | 61 | 59 (96.7%) | 8 (13.6%) | 2 (3.4%) | 3 (5.1%) | 3 (5.1%) |
|  | CRT | 27 | 26 (96.3%) | 4 (15.4%) | 2 (7.7%) | 2 (7.7%) | 0 |
|  | Both | 88 | 85 (96.6%) | 12 (14.1%) | 4 (4.7%) | 5 (5.9%) | 3 (3.5%) |
| 2019 (RT & CRT) |  | 324 | 307 (94.8%) | 71 (23.1%) | 33 (10.7%) | 31 (10.1%) | 7 (2.3%) |
| 2018 (RT & CRT) |  | 298 | 288 (96.6%) | 85 (29.5%) | 45 (15.6%) | 35 (12.2%) | 5 (1.7%) |

### Palliative radiotherapy

13 patients attended for palliative RT. Median age was 67 years (range 46-95 years). Dose and fractionation regimes delivered were 40Gy/15# (1), 20Gy/5# (6) and 8Gy/1# (6). 2 courses of 20Gy/5# schedules were discontinued earlier than planned as both patients were unable to tolerate treatment.

### Systemic anti-cancer treatments

38 patients received SACT between 1^st^ March 2020 and 23^rd^ June 2020. Median age was 56.5 year, range 37-79 years. 8 patients received induction chemotherapy, 10 patients received 1^st^ line palliative treatment and 20 received 2^nd^ line palliative treatment.

### Induction chemotherapy

3 patients with nasopharyngeal cancer received induction chemotherapy with cisplatin/5FU. 1 patient started treatment in February; receiving a 2^nd^ cycle during the pandemic then completed radical RT. A 3^rd^ cycle of induction chemotherapy and concurrent cisplatin was omitted because of concerns over COVID-19 risk. 1 patient received 2 cycles then relocated out with our cancer network. The 3^rd^ patient received 2 cycles of induction chemotherapy with radical chemoradiotherapy ongoing.

5 patients with locally advanced HNSCC received 3-4 cycles of induction chemotherapy with carboplatin/paclitaxel during this period, achieving a partial response and going on to receive radical radiotherapy.

### 1^st^ line palliative SACT

10 patients with HNC have received 1^st^ line palliative SACT during this period; carboplatin/paclitaxel (5), carboplatin/5FU (3), cisplatin/ capcitabine(1) and pembrolizumab (1).

5 patients who had commenced palliative SACT prior to the pandemic stopped treatment earlier than planned in March 2020, reasons were: toxicity (1), toxicity and concern over COVID risk (1), COVID risk alone (1), while 2 achieved a partial response after 3-4 cycles and received no further 1^st^ line treatment because of COVID-19 risk.

5 patients commenced 1^st^ line palliative SACT during the pandemic. All treatments continue to be delivered as planned.

### 2^nd^ line palliative SACT

20 patients with HNC received 2^nd^ line SACT, all with the PD-1 checkpoint inhibitor, nivolumab.

5 patients who had commenced nivolumab prior to this period discontinued treatment, 4 because of progressive disease, all have subsequently died. The other had completed a 2-year course with a complete response and remains disease free. 11 patients started nivolumab prior to the COVID-19 pandemic and have continued treatment throughout this time. 4 further patients have commenced nivolumab since March 2020.

### Palliative care

39 (19.5%) patients discussed in the regional tumour boards went on to receive palliative or supportive care only, with no active anti-cancer treatment. The median age of these patients was 71 years (range 40—91). 76.9% had multiple co-morbidities. The risk of palliative SACT in the context of COVID-19 pandemic was felt to outweigh potential benefits of treatment for 2 patients who may ordinarily have received this. For the remaining patients, the decision for palliative care was made on the basis of disease extent, performance status and co-morbidities without influence by COVID-19 risk.

## Discussion

Previous HNC specific publications have provided guidance on the modification of treatment for patients during the COVID19 pandemic. [29-33] There have been calls for increased use of hypofractionnated radical radiotherapy to mitigate the perceived risk of infection with prolonged treatment schedules, despite no evidence of increased COVID-19 infection with standard schedules, or high-quality data to support these regimens. [34]

For the first time, we report on the successful delivery of standard radical RT schedules and attenuated concurrent chemotherapy in 99 patients who received radical RT or CRT during the peak of the pandemic in the UK. While 42.6% of patients with stage III/IV HNSCC received RT alone, this was generally for reasons other than COVID-19 risk. Mean dose density of cisplatin achieved in those undergoing concurrent CRT was 151mg/m^2^. Both these findings are in keeping with pre-COVID-19 institutional data from an observational study where only 47% of our patients received cisplatin 200mg/m^2^ concurrent with RT, suggesting that eligibility for concurrent cisplatin and dose delivered in those selected have not been significantly modified on the basis of COVID-19 risk. [35]

The proportion of patients completing radical RT or CRT with gaps in treatment was 14.1%, this compares favorably with local audit data from 2018 (29.5% missed 1 or more days of RT) and 2019 (23.1%). The proportion of patients not completing radical RT or CRT during the peak of COVID-19 was 3.4%. Again, this is comparable to previous years; 2018 – 3.4% and 2019 – 5.2%. There is no suggestion therefore that patients are acquiring COVID-19 during treatment and missing, or failing to complete, treatment as a result.

Despite attending the cancer centre daily for 6 weeks, no patient contracted COVID-19 while receiving (C)RT. The 30-day mortality rate for patients treated radically with (C)RT was 2.3% (2 of 88), again comparable to previously reported outcomes in our cancer network (30-day mortality following (C) RT in 2016 – 2019 was 0.0 – 5.6%). [4-7] While it is possible that the 2 sudden deaths on treatment were due to COVID-19, both patients were generally well prior to unexpected cardiorespiratory arrest and had a history of ischaemic heart disease, making a cardiac event a more likely cause. Previous retrospective work examining the determinants of severity of COVID-19 infection in patients with cancer found those who had received SACT within 4 weeks of COVID-19 presentation were at higher risk of severe infection or death than patients not recently treated with anti-cancer therapies. [11 -13, 36, 37] Subsequent, prospective data found no significant effect on mortality for patients with COVID-19 that had received immunotherapy, targeted therapy or radiotherapy treatment in the 4 weeks prior to infection. The same was true of chemotherapy after adjusting for age, gender and co-morbidities. [38] Despite this, it is acknowledged that ‘many oncology units across the UK have eschewed palliative SACT because of concerns relating to increased risks of adverse outcomes of COVID-19’. [30] Palliative SACT was delivered to 30 patients in our cancer network at the peak of the pandemic. No cases of COVID-19 infection have been detected in these patients. Mortality has been due to progressive disease, not infection. A further 8 5 received induction chemotherapy, with no COVID-19 infection. The numbers of patients with HNC receiving SACT in our cancer centre were lower than usual. Local audit data shows on average 17 patients per week with HNC received SACT in January and February 2020. This dropped to a low of 5 patients in mid-May 2020. This reduction appears to be driven by lower numbers of patients presenting for treatment, rather than SACT being withheld in large numbers of patients because of COVID-19. This is in keeping with national data suggesting patients have delayed seeking medical help during the pandemic with a drop in the number of ‘urgent, suspected cancer ’referrals. [39]

39 patients (19.5%) were managed with supportive care only and no active anti-cancer treatment. This is in keeping with previous work in our region pre-COVID-19 which demonstrated that 21% of cases were unsuitable for active anti-cancer treatment, reflecting the significant co-morbidities, poor PS and advanced disease stage we frequently encounter in our cancer network and confirming that patients are not being denied anti-cancer treatment as a result of COVID-19. [40] To date, epidemiological studies have included patients with several different types of cancer and confirmed COVID-19 infection; patients with HNC have made up only a small proportion of these cases. In addition, it is difficult to put the potential risks of COVID-19 infection as a result of HNC treatment into perspective without knowledge of the outcomes of all patients treated during the pandemic.

This study details all patients with HNC in our cancer network managed at the peak of the COVID-19 pandemic in the UK. While some modifications have been made to our standard treatment protocols and the risks and benefits of treatment further assessed on a case-by-case basis, for the majority of patients, anti-cancer treatment has been delivered in a relatively normal fashion, albeit with altered supporting infrastructure. This strategy appears to have been successful. Only 2 of 200 (1.0%) patients have confirmed COVID-19 infection, both prior to embarking on radical RT. No patient with HNC acquired COVID-19 infection while attending for radical or palliative oncology treatment.

Our study population has multiple potential risk factors for COVID-19 infection, increased severity of infection and subsequent death – active cancer, anti-cancer treatment, other co-morbidities and social deprivation. The infection rate in the region was also significant. Yet, the rate of COVID-19 infection in this cohort of patients with HNC was low and not clinically severe.

As the first wave of the pandemic eases, attention has turned to restoring services for non COVID-19 related illness including cancer. The threat of second or subsequent waves means that the potential benefits of anti-cancer treatment still need to be balanced against the risk of COVID-19 infection. For the first time we describe encouragning early outcomes from treatment delivered during the peak of the pandemic in patients with HNC. We hope that our data provides some reassurance to clinicians and patients with HNC in these challenging times.

## Conclusion

It is feasible and safe to deliver meaningful treatment for patients with HNC during the COVID-19 pandemic for a high-risk population in a region with a significant rate of infection.

## Data Availability

Data will be made available on request

## Acknowledgments

We are grateful to Eva Stalker, MDT Coordinator, Queen Elizabeth Hospital, Glasgow for tumour board data and to Christine Crearie, West of Scotland CEPAS clinical support team, senior information analyst for SACT data.

